# Imputed Gene Expression versus Single Nucleotide Polymorphism in Predicting Gray Matter Phenotypes

**DOI:** 10.1101/2023.05.05.23289592

**Authors:** Jiayu Chen, Zening Fu, Armin Iraji, Vince D. Calhoun, Jingyu Liu

## Abstract

Genetics plays an important role in psychiatric disorders. A clinically relevant question is whether we can predict psychiatric traits from genetics, which holds promise for early detection and tailored intervention. Imputed gene expression, also known as genetically-regulated expression (GRE), reflects the tissue-specific regulatory effects of multiple single nucleotide polymorphisms (SNPs) on genes. In this work, we explored the utility of GRE for trait association studies and how the GRE-based polygenic risk score (gPRS) compared with SNP-based PRS (sPRS) in predicting psychiatric traits. A total of 13 schizophrenia-related gray matter networks identified in another study served as the target brain phenotypes for assessing genetic associations and prediction accuracies in 34,149 individuals from the UK Biobank cohort. GRE was computed leveraging MetaXcan and GTEx tools for 56,348 genes across 13 available brain tissues. We then estimated the effects of individual SNPs and genes separately on each tested brain phenotype in the training set. The effect sizes were then used to compute gPRS and sPRS in the testing set, whose correlations with the brain phenotypes were used to assess the prediction accuracy. The results showed that, with the testing sample size set to 1,138, for training sample sizes from 1,138 up to 33,011, overall both gPRS and sPRS successfully predicted the brain phenotypes with significant correlations observed in the testing set, and higher accuracies noted for larger training sets. In addition, gPRS outperformed sPRS by showing significantly higher prediction accuracies across 13 brain phenotypes, with greater improvement noted for training sample sizes below ∼15,000. These findings support that GRE may serve as the primary genetic variable in brain phenotype association and prediction studies. Future imaging genetic studies may consider GRE as an option depending on the available sample size.

## INTRODUCTION

Genetics is known to play an important role in psychiatric disorders^1-3^. Genetic influence is reflected in increased family risk and has been further quantified through family studies ^4-6^. A recent study leveraging the Swedish sibling cohort conducted a comprehensive investigation on eight psychiatric traits and the reported heritability varied from 0.3 for major depressive disorder to 0.8 for attention-deficit/hyperactivity disorder, with an estimate of 0.6 for schizophrenia^7^. The significant progress in large-scale genome-wide association studies (GWASs) in the past decade provides more direct evidence for genetic effects on psychiatric disorders. Starting from the Psychiatric Genomic Consortium (PGC) GWAS of schizophrenia (SZ) as a milestone in 2014, GWASs started to yield reliable and generalizable risk single nucleotide polymorphisms (SNPs) for complex psychiatric traits of high polygenicity^8-14^ with sample sizes exceeding 100K. These GWASs have not only revealed high-risk genomic loci, but also grounded new approaches of heritability estimation leveraging the genomic profiles^15^. The GWAS-based heritability estimates are usually lower than those from family studies but show significant correlations^7^.

At this point, a more clinically relevant question is whether we can predict a specific psychiatric trait from genetics, which holds promise for early detection and tailored intervention. Given the high polygenicity, the risk SNPs identified by GWASs in general show modest effect sizes and lack predictive power at the univariate level. This has motivated the polygenic risk score (PRS) approach that is to aggregate the effects of multiple SNPs with each SNP weighted by the effect size estimated from reliable GWASs. As a multivariate measure, PRS has shown improved power, e.g., roughly explaining 11% of the variance in liability for schizophrenia and 4% for bipolar disorder ^9,11,16^. However, computing PRS requires a reliable GWAS with a decent sample size as a prerequisite, which is not always readily available. For instance, many behavioral and cognitive measures are more difficult to harmonize across cohorts^17,18^, making data aggregation more challenging. And GWASs of brain phenotypes are also lagging in terms of sample size^19-22^.

In parallel, there is another line of effort to integrate functional annotations with GWASs to improve statistical power and interpretability. One typical example is imputed gene expression, also known as genetically regulated expression (GREs)^23^. GREs are grounded by the observation that a subset of genomic SNPs regulate gene expression, so-called expression quantitative trait loci (eQTLs). Thus, leveraging public resources such as the Genotype-Tissue Expression (GTEx) project^24^, models can be constructed to characterize the relationships between the expression level of a specific gene and its tissue-specific eQTLs. This allows GREs to be imputed for any individuals provided that their genotypes are available^23,25^. And the gene-eQTL relationship can be combined with SNP-based GWAS summary statistics to estimate the effects of imputable genes on the traits that have been investigated in the GWASs, known as transcriptome-wide association studies (TWAS)^26^. Like PRS, TWAS also needs GWAS as a prerequisite, which is a limiting factor.

An intriguing question is whether it’s feasible to conduct trait association studies using GREs as the primary genetic variable and how the GRE-based risk score (denoted as gPRS in the following text) compares with SNP-based PRS (denoted as sPRS) in predictive utility. For each imputable gene, the GRE combines the effects of multiple eQTLs, so theoretically we expect them to carry larger effect sizes than SNPs and boost statistical power in association tests. Indeed this is the motivation for proposing GRE, and finds support from the TWAS results^26^. With that said, one recent study computed gene expression-based risk scores using GREs weighted by TWAS effect sizes, which however did not outperform sPRS in predictive power at a sample size level of 50K^27^. There are two points here that deserve further investigation. First, rather than directly assessing the effects of genes, TWAS builds up gene-trait associations based on SNP-trait associations of available GWASs by integrating eQTL information. It remains unclear if we can circumvent GWAS to directly use GREs for trait association analyses and still obtain a gain in statistical power. Second, a more comprehensive investigation is needed to compare gPRS with sPRS in terms of predictive power for different levels of sample sizes. A possible scenario is that the gain of using GREs may be more substantial for smaller sample sizes, where SNPs associations are more vulnerable to the power issue. And when the sample size becomes sufficiently large, sPRS is expected to outperform gPRS as the latter only leverages a portion of the genome (i.e., eQTLs). Considering that a sample size of 50K is not always accessible, it is important to examine how the performance varies with sample sizes, to inform future experimental designs.

The current work aims to assess the applicability of GREs in directly assessing gene-brain phenotype associations and the performance of the resulting gPRS in predictive brain phenotypes derived noninvasively from magnetic resonance imaging (MRI), particularly how it compares with sPRS across a range of sample sizes. We are particularly interested in brain phenotypes, as compared to GWASs of psychiatric disorders, understanding genetic contributors to brain abnormalities better informs the pathology. However, even with data aggregation, it is still difficult to reach a large sample size in GWASs of brain phenotypes, largely due to the availability of both imaging and genomic data. This has motivated the exploration of GREs for improved power with limited samples. In addition, GREs promise, to a certain level, tissue-specificity of genetic correlates underlying brain phenotypes, which is a tempting scenario given the difficulty in obtaining brain tissues. Specifically in this study, we conducted training and testing using the UK Biobank (UKB) data and compared the predictive power of gPRS and sPRS on SZ-related gray matter phenotypes derived from structural magnetic resonance imaging (sMRI) data.

## MATERIALS AND METHODS

### UK Biobank

The current work leveraged the population-based UKB cohort which recruited more than 500K individuals across the United Kingdom. The UKB study was approved by the North West Haydock Research Ethics Committee, and the data used in our work were obtained under data application number 34175. Specifically, we used the imputed SNP data and T1-weighted MRI data of 34,149 European ancestry individuals with both modalities available after quality control (QC), including 16,063 males and 18,086 females, aged between 45-81 with a median of 64 when brain MRI was collected.

### Genetic data

The imputed SNP data released by UKB consisted of 487,320 individuals and ∼96 million variants (v3_s487320). Details of genotyping and imputation can be found in the paper that describes the UKB genomic data^28^. In brief, DNA was extracted from blood samples and genotyping was carried out by Affymetrix Research Services Laboratory. Most of the individuals (94%) reported their ethnic background as ‘white’, which was a broad-level group. SHAPEIT3 was used for phasing^29^. Imputation was conducted using IMPUTE4^28^ with the Haplotype Reference Consortium reference panel^30^ and the UK10K Consortium reference panel^31^.

In this study, we first identified the participants that passed the UKB quality control (sex mismatch, missing rate, and heterozygosity) and also had sMRI data available. We then excluded SNPs with minor allele frequencies < 0.01, as recommended for subsequent GWAS and PRS analysis^32^. We then conducted relatedness estimation (identify-by-descent) using PLINK^33^. For each group of individuals that were second-degree relatives or closer, only one individual was retained for subsequent analysis. Finally we identified individuals of European ancestry (in a more strict sense) to be those close (< 3SD) to the center of the ‘white’ cluster as defined by the first four principal components.

### Structural MRI data

The UKB imaging enhancement plan starting in 2014 highlights the aim of reinviting 100K participants for multi-modal imaging^34^. The data we used contain T1 scans of ∼37K individuals. UKB used identical scanner models, coils, software, and protocols across centers to ensure data harmonization as much as possible. The T1 scans used the magnetization-prepared rapid acquisition with gradient echo sequence, resolution = 1.0 mm × 1.0 mm × 1.0 mm, matrix = 256 × 256 × 208, TI = 880 ms, TR = 2000 ms, parallel imaging acceleration factor = 2.

The whole brain T1-weighted data were preprocessed using the standard statistical parametric mapping 12 voxel-based morphometry pipeline as described in our previous work^35,36^. With the unified model integrating image registration, bias correction, and tissue classification, the resulting gray matter volume (GMV) images were estimated by the modulated method and then resliced to 1.5 mm × 1.5 mm × 1.5 mm. The resliced GMV images were further smoothed by a 6 mm full width at half-maximum Gaussian kernel. We calculated the gray matter mask based on the average GMV > 0.2, which included 416,407 gray matter voxels for further analyses. We excluded 55 outlier participants whose GMV profiles (masked) showed correlations < 0.8 with the average GMV profile across all the participants.

### Imputation of gene expression

MetaXcan was used to impute gene expression from genotypes^25^ leveraging the GTEx V8 release^24^ for eQTL information. Given the goal to evaluate the predictive power of gPRS for brain phenotypes, we chose to focus on the GREs of brain tissues. The imputed SNPs that passed the aforementioned quality control (QC) were used as input. As a result, a total of 56,348 genes were successfully imputed across 13 available brain tissues. **Figure 1a** shows a breakdown of the number of imputed genes for individual brain tissues.

**Figure 1:**
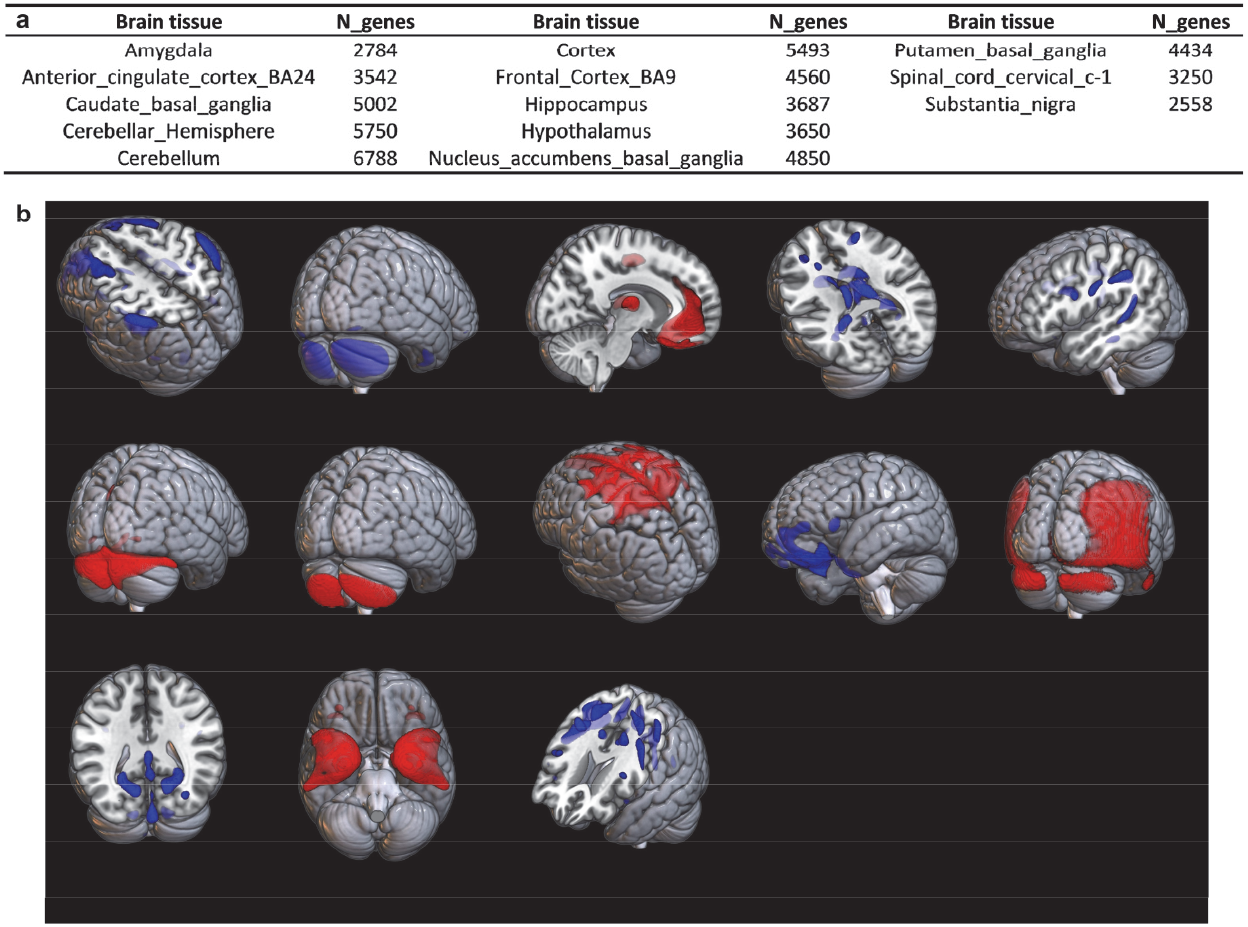
(a) A summary of imputed genes for each of the 13 brain tissues; (b) Spatial maps of the 13 schizophrenia-related brain networks.

### Gray matter phenotypes

This work investigated a total of 13 SZ-related brain networks for predictability by genetics. These brain networks were derived by applying independent component analysis (ICA)^37,38^ to GMV data from other studies (COBRE, FBIRN, and BSNIP^36,39^), called source-based morphometry^40^. ICA decomposes the GMV data into a linear combination of maximally spatial-independent components. Each component or brain network essentially identifies a pattern of voxels with covarying GMV patterns, and the component’s associated loadings reflect how this brain network is weighted or expressed in different subjects. For these 13 brain networks, their loadings have been found to show robust SZ relevance across cohorts of different age ranges, including significant group differences between controls and individuals with SZ in adults, significant associations with Structured Clinical Interview for DSM-5 (SCID) SZ scale in young adults, as well as significant associations with prodromal psychosis scale in adolescents (unpublished data). **Figure 1b** shows the spatial maps of these 13 SZ-related GMV networks, where the red and blue colormaps indicate that the original voxel-level GMVs were positively or negatively correlated with the component loadings extracted by ICA, or the brain phenotypes tested in the current work. For all these highlighted brain regions, cases with SZ showed lower GMV compared to controls.

We conducted spatially-constrained ICA, an sMRI version of our fully automated NeuroMark pipeline^41^, on the UKB GMV data with the 13 SZ-related brain networks serving as references. This pipeline allowed obtaining brain networks for the UKB individuals that corresponded to the reference networks while allowing variations specific to the UKB data. Similarly, this pipeline yielded associated loadings of components that reflected the weights of brain networks on subjects, which were used as gray matter phenotypes to be predicted by GREs.

## SNP-based polygenic risk score

First we made sure that the QC was calibrated with those recommended for GWAS and PRS analysis^32,42^. We then chose to prune the SNP data before running GWAS, rather than conducting GWAS on unpruned data followed by clumping + thresholding, mainly to reduce computation burden^32,42^. The QC‘d SNPs went through a heavy pruning (r^2^ < 0.1, 500 kb window) resulting in 208,752 autosomal SNPs. GWAS was conducted on the training data using PLINK to assess the additive effects of individual SNPs on one brain phenotype (continuous variable) at a time. Age, sex, MRI scanning site, as well as the top 5 principal components of the genomic SNP data were included as covariates. Then in the testing data sPRS represented the combined effects of SNPs that passed the specified p-value threshold. In this work, we tested five p-value thresholds including 0.0001, 0.05, 0.1, 0.5, and 1.

### Gene-based polygenic risk score (gPRS)

The GREs of imputed 56,348 genes across 13 brain tissues were assessed for associations with each brain phenotype using a regression model in the training data, controlling for age, sex, MRI scanning site, and the top 5 principal components for population structure. The previous work suggested that in general, the full model (p-value threshold = 1) multi-tissue gene risk scores showed stronger predictive power^27^. Consequently, in this work gPRS represented the combined effect of all the GREs from 13 brain tissues.

It is not well established whether pruning is needed for gene-based risk scores, given that each gene has its own biological function and impact on the traits. With that said, to address the concern of including highly correlated genes might inflate the associations between the resulting gPRS and gray matter phenotypes, we also examined how gPRS with pruning compared with sPRS in prediction. Specifically, we conducted a 500K-window pruning with r^2^ < 0.16 on all the 56,348 imputed genes across 13 brain tissues. After pruning, a total of 16,527 tissue-specific gene markers were retained for gPRS analysis, denoted as gPRS_pruned in the following text.

### Training and testing

The UKB data were partitioned into 30 folds to examine the impact of training sample size on the predictive power of sPRS and gPRS. For each random partition, one fold of 1,138 individuals was used for testing, while the training sample size increased from one fold (N = 1138) up to 29 folds (N = 33,011). For each training set, we estimated the effects of individual SNPs and genes on brain phenotypes as described above. The resulting summary statistics were then used to compute sPRS and gPRS in the hold-out testing set (i.e., predicted brain phenotypes). The correlations (R) between the predicted and observed brain phenotypes were employed as a metric of prediction accuracy. A total of five random partitions were conducted to characterize how the overall predictive power for all the brain phenotypes varied with training sample size, and if there were any statistical differences in the predictive power between sPRS and gPRS.

### Heritability

We also examined how heritability might impact sPRS and gPRS performance. We conducted GWAS on each brain phenotype using the common SNPs of the QC’ed UKB data and the HapMap SNPs, as recommended by the LD Score (LDSC) regression tool ^15^. The full UKB data covering 34,149 European ancestry individuals were included in the GWAS for heritability analysis. Additive effects were evaluated for individual SNPs, controlling for the same covariates as described above. Finally, heritability was estimated for each brain phenotype based on the summary statistics.

## RESULTS

**Figure 2a** shows how prediction accuracies varied across the tested range of training sample sizes for both gPRS and sRPS, where five different p-value thresholds were explored for sPRS from 0.0001 up to 1. **Figure 2b** shows the prediction accuracies of gPRS_pruned compared with gPRS and sPRS. Each data point and its confidence interval reflect the mean and standard error of the observed prediction accuracies across 13 brain phenotypes and 5 random partitions for a specific training sample size. It can be seen that, overall, sPRS prediction accuracies improved with higher p-value thresholds and more SNPs included. This performance improvement saturated around the p-value threshold of 0.5, given that the prediction accuracies didn’t differ significantly between the p-value threshold of 0.5 and 1. We then focused on sPRS with a p-value threshold of 1 for the primary comparison with gPRS.

**Figure 2:**
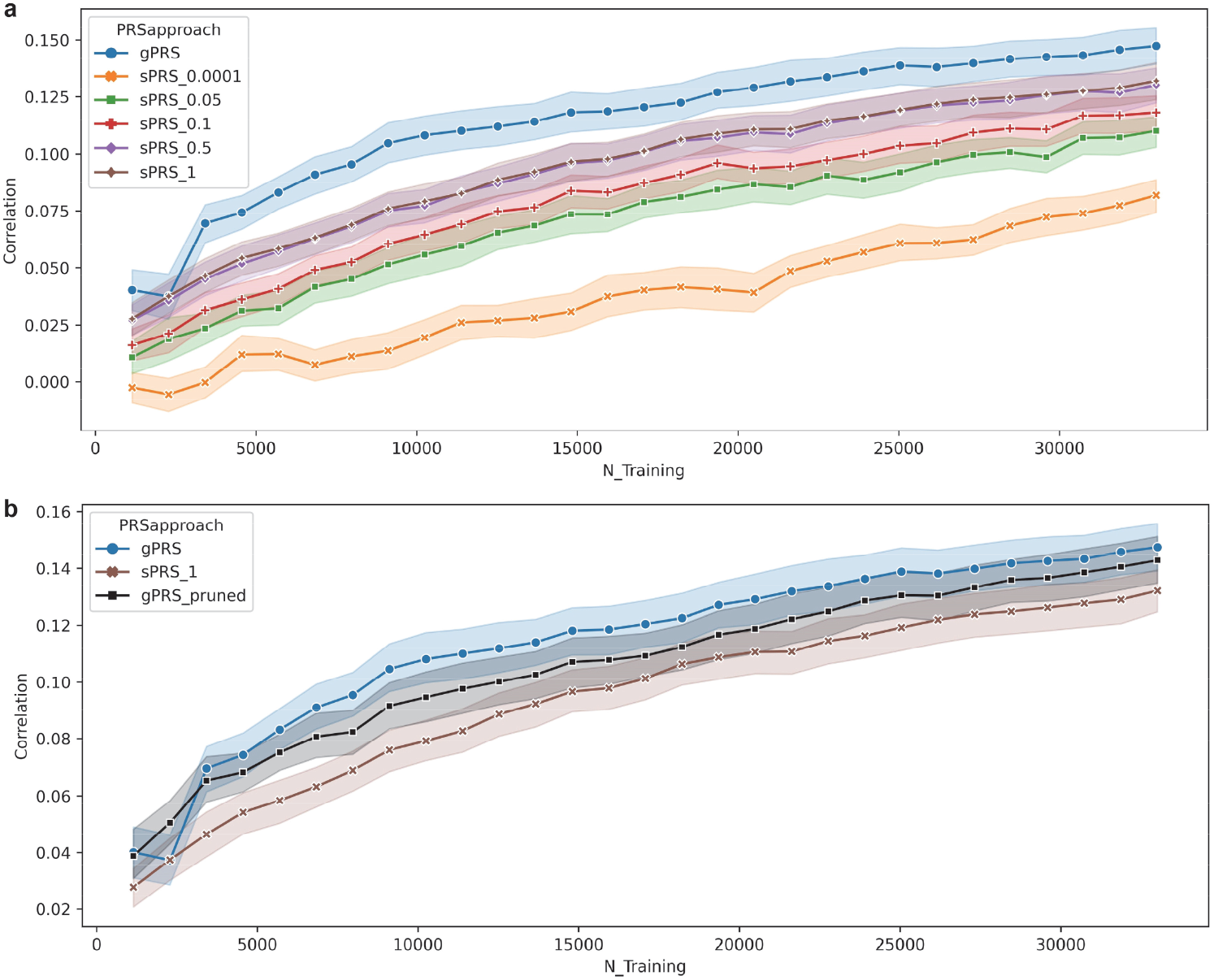
(a): Changes in prediction accuracies with increasing training sample sizes. Each data point reflects the mean and standard error of the prediction accuracies observed from all 13 brain phenotypes across 5 random partitions at a specific training sample size. For the sPRS approach, the results of all the tested p-value thresholds from 0.0001 up to 1 are provided. (b): Prediction accuracies of gPRS_pruned compared with gPRS and sPRS.

Both gPRS and sPRS showed improved prediction accuracies with increasing training sample sizes. Starting from a training sample size of ∼1,100, the mean accuracy of gPRS increased from ∼0.04 up to ∼0.14 at a training sample size of ∼33K. In parallel, the mean prediction accuracy of sPRS increased from ∼0.024 to 0.11. Notably, the training sample size was capped at ∼33K in the current study, and the increasing trend of prediction accuracies hadn’t shown any obvious sign of saturation. With pruning, gPRS_pruned still outperformed sPRS, although showing lower accuracies compared to gPRS.

Figure 3. shows the side-by-side violin plots of gPRS and sPRS predictive accuracies for all the tested training sample sizes. For each sample size, we conducted a two-sample t-test based on the prediction accuracies of 13 gray matter phenotypes and 5 random partitions, to examine whether gPRS outperformed sPRS. Detailed statistics are also included, where a positive t-value indicates gPRS showing a higher mean prediction accuracy compared to sPRS. For almost all the tested sample sizes, gPRS showed significantly higher prediction accuracies than sPRS, with p-values ranging from 0.04 to the lowest 1.37E-06 which was observed at a training sample size of 6,828. The only exception was that no significant difference was noted for a training sample size of 2,276.

**Figure 3:**
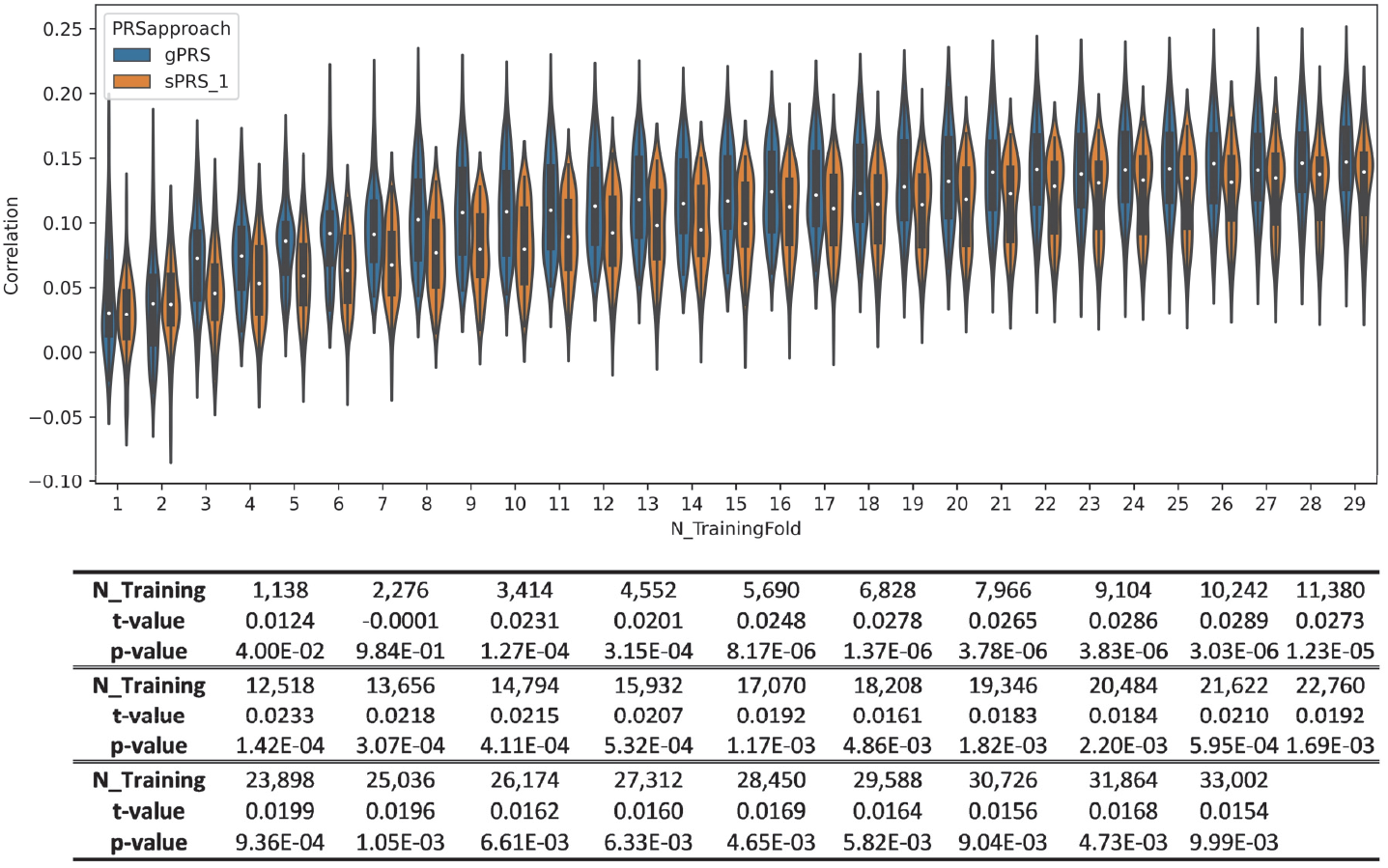
Comparison of prediction accuracies between gPRS and sPRS across the whole tested range of training sample sizes. For each training sample size, the violin plots show the distributions of prediction accuracies of 13 brain phenotypes across 5 random partitions of gPRS and sPRS respectively. The table below summarizes the t-values and p-values of the two-sample t-tests.

We also examined how heritability might impact the gPRS and sPRS prediction accuracies. **Figure 4** shows how prediction accuracies varied across brain networks showing different levels of heritability. Each data point indicates the mean prediction accuracy (along with the confidence interval) across all the training sample sizes for one specific brain phenotype, sorted by heritability. All the brain phenotypes were significantly heritable, with estimated h^2^ ranging from 0.28 to 0.35. Meanwhile, no significant association was observed between the estimated heritability and the sPRS/gPRS prediction performance. Despite discrepancies noted for 2 out of 13 brain phenotypes, the predictability was consistent between sPRS and gPRS for the remaining brain phenotypes, where a higher gPRS accuracy was accompanied by a higher sPRS accuracy. And gPRS showed improved prediction accuracies for most of the brain phenotypes compared to sPRS, while comparable accuracies were noted for the rest.

**Figure 4:**
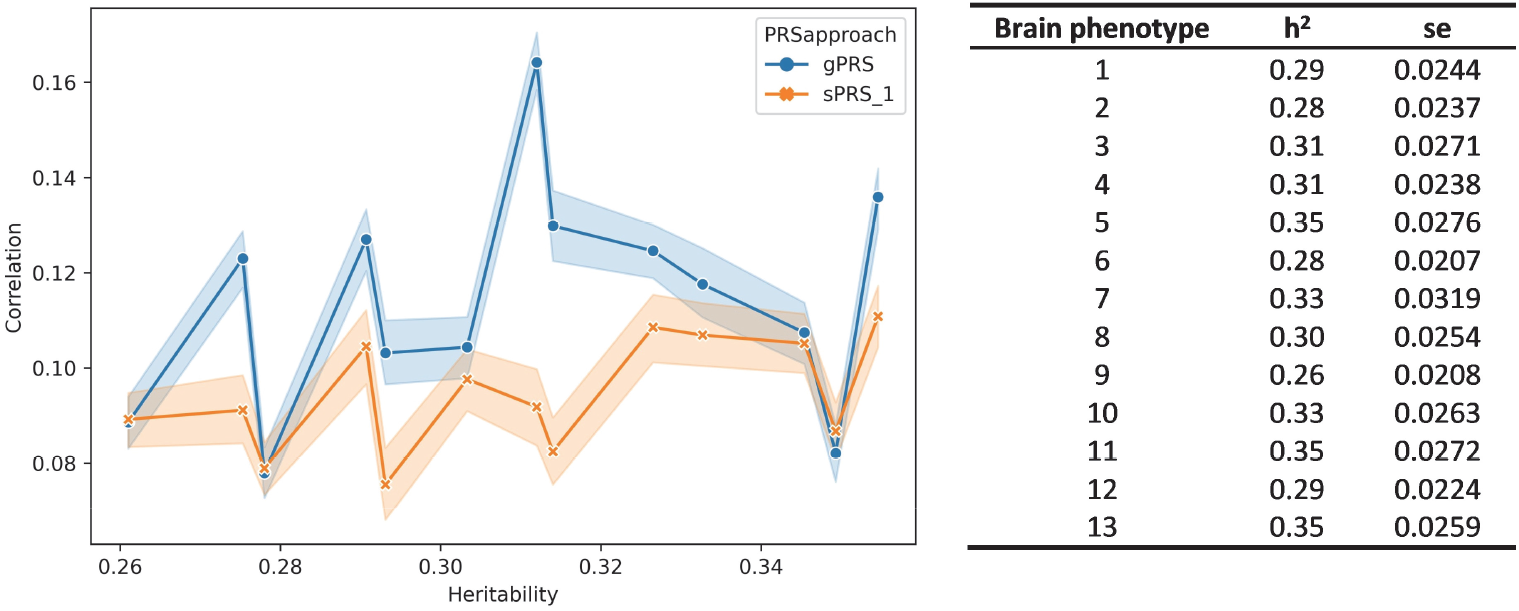
Scatter plots of prediction accuracies versus estimated heritability of the schizophrenia-related brain networks. Each data point reflects the mean and standard error of the prediction accuracies observed from all the training sample sizes across 5 random partitions for a specific brain phenotype.

## DISCUSSION

We investigated the applicability of GREs for directly assessing gene-brain phenotypes associations and gPRS prediction. A total of 13 SZ-related gray matter networks were employed as brain phenotypes. We partitioned the UKB data into training and testing sets to compare the predictive power of gPRS and sPRS on these brain phenotypes, and how the predictive power varied across a wide range of training sample sizes.

As expected, both sPRS and gPRS showed improved predictive power with increasing training sample sizes. Also, both curves showed sharper increases at smaller sample sizes in which increasing the training samples by 1,000 yielded more gain in prediction accuracy compared to a sample size increase from 20,000 to 21,000. With that said, the prediction accuracy curve did not quite saturate at the largest tested sample size of 33K. It remains a question what the most cost-effective sample size should be for this type of risk score study of brain phenotypes, which also depends on the trait heritability, polygenicity, risk prevalence, etc.

The gPRS computed based on GREs reliably predicted gray matter phenotypes, even at small training sample sizes. With only 1,138 training samples, the gPRS prediction accuracies observed across 13 gray matter networks and 5 random partitions were significantly greater than 0. It should be emphasized that the computation of gPRS didn’t leverage any large-scale GWAS. The results support that GREs can be used as primary genetic variables for directly assessing gene-brain phenotype associations and can yield generalizable results with a reasonable sample size.

It’s noted that gPRS, either without or with pruning, outperformed sPRS in predictive power for almost the whole tested range of training sample sizes. The main comparison as shown in Figures 2 and 3 was based on the prediction accuracies across all the 13 networks and 5 random partitions, which was expected to better characterize the overall predictive power and how it related to the training sample size. Furthermore, Figure 4 shows the gPRS and sPRS prediction accuracies of individual gray matter networks, where the improvement in gPRS prediction was relatively evenly distributed across all the 13 networks, rather than being majorly driven by one brain network. These findings lend support that gPRS presents improved prediction accuracies on brain phenotypes in a general sense, which does not appear to be specific to certain brain regions. More significant improvement in prediction was noted for a sample size range of ∼3000-15000, where using gPRS improved the mean prediction accuracy by 0.019-0.029. At larger sample sizes, the sPRS performance was more comparable to gPRS, although gPRS remained to show significantly higher accuracies up to 33K. This observation echoes the speculation that imputed genes, as a multivariate factor of SNPs, alleviate the issue of modest effect sizes and the gain of using GREs is more substantial for smaller sample sizes. Meanwhile, larger sample sizes above 33K need to be tested to locate the range where sPRS outperforms gPRS by leveraging the whole genome rather than just eQTLs. Collectively, these findings promote GREs over SNPs for brain phenotype association and prediction analysis with a sample size below 30K.

One speculation is that the improved prediction noted in gPRS might be related to tissue-specificity. Only imputed genes of brain tissues were used to compute gPRS, which was expected to align better with the target traits of brain phenotypes. While GREs reflect multivariate regulatory effects of SNPs, using only imputed genes of brain tissues is equivalent to applying a screening on SNPs, such that only SNPs known to regulate gene expression levels in those brain tissues were included for assessment. Excluding SNPs that are less likely to directly impact the brain may help reduce the background noise in gPRS, which is expected to benefit more when the sample size is low. This is also consistent with the notion that independent filtering help boost detection power for high-throughput experiments^43^.

Both sPRS and gPRS reflect the genetic effects on the traits. As a result, the resulting prediction accuracies are expected to be capped by heritability. In the current work, all the tested brain phenotypes showed significant heritabilities, justifying their predictability by genetics. Meanwhile, no significant association was noted between heritability and prediction accuracies, which might be due to the observed heritabilities varied in a relatively narrow range of 0.28-0.35, where the impact on prediction might not be reliably captured across 13 gray matter traits. Besides, the observed prediction accuracies were well below the theoretical upper limit as indicated by heritability, suggesting that the linear model of weighted sum might not capture all the genetic effects on the traits, and more sophisticated models are needed to further boost the predictive power.

The current work should be interpreted in light of the following limitations. First, we did not test all the PRS approaches available for SNPs, some of which such as PRS with adaptive shrinkage had been reported to show improved association and prediction power compared to PRS with a simple pruning. Given that not many as sophisticated approaches were available for genes, we chose to compare gPRS with sPRS yielded by a comparable simple model. It remains to be elucidated whether gPRS performance could also benefit from more advanced learning processes as sPRS did. Second, we did not optimize the p-value threshold of sPRS for individual training and testing sets using nested cross-validation. Meanwhile, we did test five p-value thresholds from 0.0001 to 1, and the results consistently indicated that a higher p-value threshold with more SNPs included for sPRS overall yielded improved prediction accuracies on the 13 gray matter phenotypes across all the tested training sample sizes. A more detailed breakdown of how prediction accuracies changed with p-value thresholds for individual brain phenotypes and random partitions (Figure S1) further confirmed the consistency in performance improvement. These observations suggest that although not finely tuned for optimal prediction in each test, the reported main results of sPRS with a p-value threshold of 1 are not expected to be dramatically poorer than the optimal accuracy. And this should not impact the comparison between sPRS and gPRS, given the latter was not test-optimized either. Third, the training sample size was capped at ∼33K, where we hadn’t seen a turning point where sPRS started to outperform gPRS. This needs to be explored further to better inform future experiment designs. Fourth, the current work only assessed gPRS and sPRS performances on 13 SZ-related gray matter networks. We still need to answer whether the observations generalize to other imaging modalities, such as functional and diffusion MRI measures, and other behavioral and cognitive measures, as well as clinical assessments. Fifth, while PRS is a linear model, it deserves further investigation whether GREs also promise improved power in nonlinear models such as deep neural networks.

In summary, we provide evidence that GREs hold promise for serving as the primary genetic variable in brain phenotype association and prediction studies, which are likely more powered than SNPs, particularly when the sample size is relatively small (< 15K). Future imaging genetic studies may consider GREs as an option depending on the available sample size.

## Data Availability

All data produced in the present study are available upon reasonable request to the authors.

## ACKNOWLEDGEMENTS

This project was funded by the National Institutes of Health (P20GM103472, R01EB005846, 1R01EB006841, R01MH106655, 5R01MH094524) and the National Science Foundation (1539067).

## Supplemental Figure

**Figure S1:**
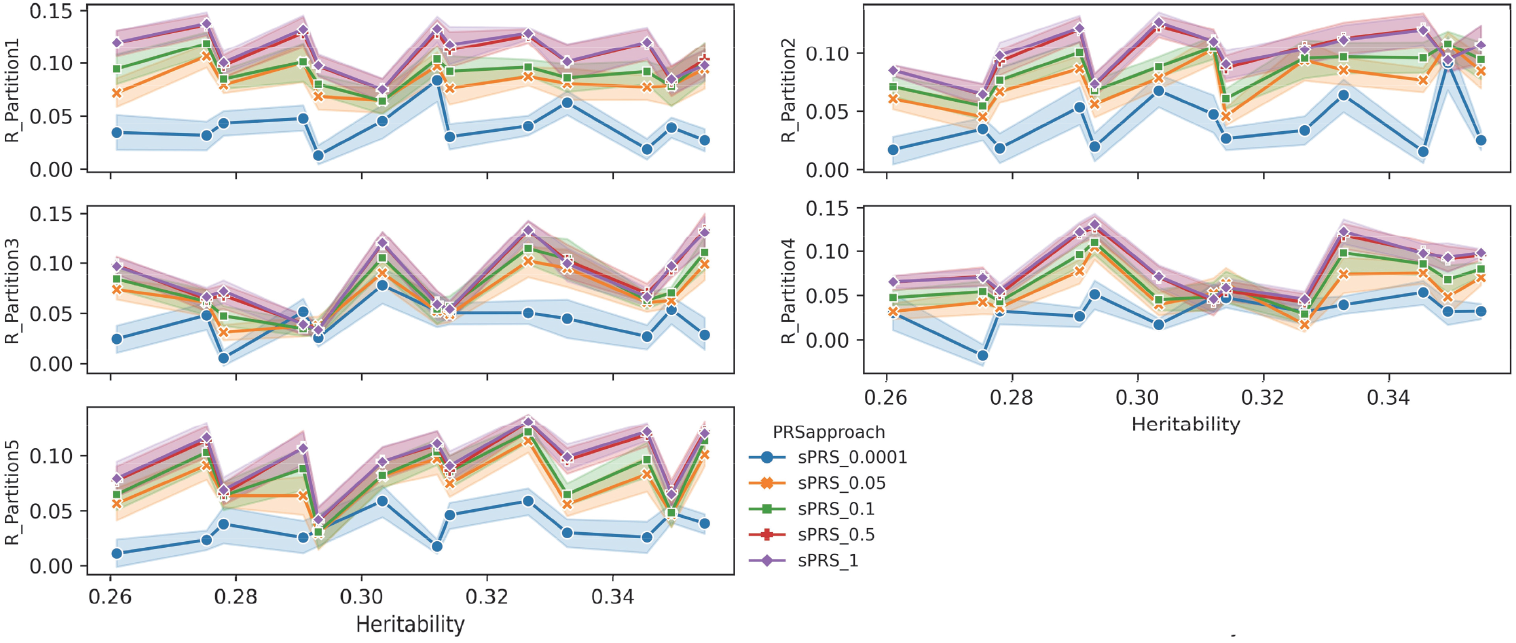
Prediction accuracies observed across all the training sample sizes for individual brain phenotypes and individual random partitions. Each subplot shows the performance of one random partition, where each data point represents a specific brain phenotype and shows the mean and standard error of the prediction accuracies across 29 tested training sample sizes.

